# Explainable AI in Deep Learning-based Detection of Aortic Elongation on Chest X-ray Images

**DOI:** 10.1101/2023.08.28.23294735

**Authors:** Estela Ribeiro, Diego A. C. Cardenas, Felipe M. Dias, Jose E. Krieger, Marco A. Gutierrez

**Author notes:** **Correspondence to:** Estela Ribeiro.

## Abstract

**Aim:** Aortic Elongation can result from age-related changes, congenital factors, aneurysms, or conditions affecting blood vessel elasticity. It is associated with cardiovascular diseases and severe complications like aortic aneurysms and dissection. We aim to assess qualitatively and quantitatively explainable methods in order to understand the decisions of a deep learning model for Aortic Elongation detection with Chest X-Ray (CXR) images.

**Methods and Results:** In this work, we evaluated the performance of deep learning models (DenseNet and EfficientNet) for aortic elongation detection based on transfer learning and fine-tunning techniques using CXR as input. DenseNet achieved higher accuracy (84.7% ± 0.7), precision (75.6% ± 1.3), sensitivity (88.7% ± 2.7), specificity (82.3% ± 1.6), F1-score (81.6% ± 1.0), and AUROC (93.1% ± 0.4) than EfficientNet. To gain insights into the decision-making process of the deep learning models, we employed Grad-CAM and LIME explainability methods. Through these techniques, we were able to successfully identify the expected location of aortic elongation in the x-ray images. Moreover, we used the pixel-flipping method to assess quantitatively the interpretations providing valuable insights into models behavior.

**Conclusion:** Our study presents a comprehensive strategy for analyzing CXR by integrating Aortic Elongation detection models with explainable methods. By incorporating explainable AI techniques, we enhanced the interpretability and understanding of the models’ decisions. This approach holds promise for aiding clinicians in timely and accurate diagnosis, potentially improving patient outcomes in clinical practice.

## 1 Introduction

Aortic Elongation is a medical condition in which the aorta, the largest artery in the human body, is longer than normal [1]. This can occur for various reasons, including age-associated changes [2], congenital factors [10, 3, 6, 15, 5], or underlying medical conditions that affect the elasticity of blood vessels, resulting in Cardiovascular Diseases (CVD). Common diseases associated to aortic elongation include aortic aneurysms [9], Marfan Syndrome [10], Ehlers-Danlos Syndrome [3], Loeys-Dietz Syndrome [15] and Turner Syndrome [5]. The consequences of an elongated aorta can vary depending on the severity and cause of the condition. In some cases, an elongated aorta may not present symptoms or significant problems. In other cases, however, it can lead to serious complications such as aortic aneurysms, aortic dissection, aortic rupture, and other cardiovascular conditions [1].

The detection of aortic elongation can be achieved through various imaging examinations, including Chest X-Rays (CXR), Computerized Tomography (CT), or Magnetic Resonance Imaging (MRI). Furthermore, advancements in the Deep Learning (DL) field have significantly enhanced efforts aimed at automating disease detection using these imaging modalities. DL offers a robust set of tools for processing and analyzing medical images, thereby greatly benefiting research focused on the automatic detection of diseases.

Convolutional Neural Networks (CNNs) [19, 30], a type of DL model, are commonly used in computer vision tasks such as image and video recognition. CNNs are able to extract features from raw data without any prior knowledge of the input data. This is particularly useful for image recognition tasks where traditional feature extraction methods may not be effective. CNNs also learn multiple layers of abstraction that represent features at increasing levels of complexity. This allows them to recognize complex patterns and objects. Furthermore, they have the ability to recognize patterns that may be difficult for the human eye to notice by learning from large amounts of data. Overall, DL has the potential to greatly improve the accuracy and efficiency of medical imaging, leading to better patient outcomes and more effective healthcare.

However, it is important to carefully validate and interpret the results of DL models, as they can be sensitive to biases in the training data and may not always generalize to new patient populations [24]. Moreover, the black-box nature of the DL models prevents transparency in their decision-making processes, despite the fact that they are capable of achieving high classification accuracy [16]. Given this lack of interpretability, the applications of AI systems in hospitals may be limited [12]. This issue has gained greater attention recently, raising questions about whether AI systems should be used in high-risk situations like medicine [24, 8]. Therefore, many researchers are focusing on developing techniques that could help to understand how DL models reach their decisions, known as eXplainable Artificial Intelligence (XAI) [29, 28, 22, 14]. Some of these techniques rely on the analysis of gradients [28, 29], which can be difficult to interpret, while others are based on perturbation [22, 14], which can be specific to a particular instance.

Despite this interpretability challenge, the integration of DL models with medical knowledge presents promising opportunities, particularly in areas like radiology. Since CXR exams are quick and inexpensive, its usage for the detection of Aortic Elongation is important, given that the determination of this condition can give indicatives of a set of diseases, as aforementioned. Low-income hospitals that might not have access to experienced and knowledgeable radiologists might lessen the burden on their medical infrastructure by implementing an automatic detection model for this condition. Furthermore, even the most experienced expert may be susceptible to errors, thus such a DL model can assist in handling the arduous and time-consuming task of interpreting and evaluating CXR images. By quickening the diagnostic process and lessening the workload of doctors, the integration of DL models and medical knowledge can enhance the performance of medical staff and could also decrease patient waiting times.

In this study, we implemented two DL models to detect Aortic Elongation using CXR images obtained from a public available dataset and a private dataset using a transfer learning and fine-tunning approach. To explain our models predictions we used Grad-CAM and LIME, two widely-known explainable AI methods, to visualize which parts of the CXR are most important when our models predict aortic elongation. Additionally, we used the metric pixel-flipping to quantitatively evaluate the explainable methods, providing qualitative and quantitative insights on the models predictions. Our paper offers the following key contributions:

1. Development of DL models utilizing transfer learning and fine-tuning techniques to achieve accurate identification of Aortic Elongation in CXR images;
2. Implementation of Grad-CAM and LIME explainability methods to gain insights into the decision-making process of the DL models;
3. Comparative analysis of two distinct DL models based on their performance metrics and explainability outputs;
4. Utilization of the pixel-flipping quantitative metric to evaluate the interpretability and reliability of the explanations provided by XAI methods.

## 2 Methods

The general structure of the proposed method is shown in Figure 1. In summary, our approach involved utilizing two CNNs, namely DenseNet and EfficientNet. We initiated the training process by leveraging the ImageNet weights and then we employed transfer learning using a publicly available CXR dataset to enable our models to learn the new domain. Subsequently, we performed fine-tuning using a private CXR dataset. To assess the performance of our models, we implemented a 5-fold cross-validation strategy. Once we gained insights into their performance, we proceeded to retrain the models using the entire dataset, without employing the cross-validation process. This allowed us to create comprehensive models that were subsequently utilized for the proposed explainable AI methods. Finally, we conducted a visual analysis of the interpretations provided by the explainable methods, in conjunction with a quantitative analysis using the pixel-flipping method, on a subset of the test set from our private dataset.

**Figure 1:**
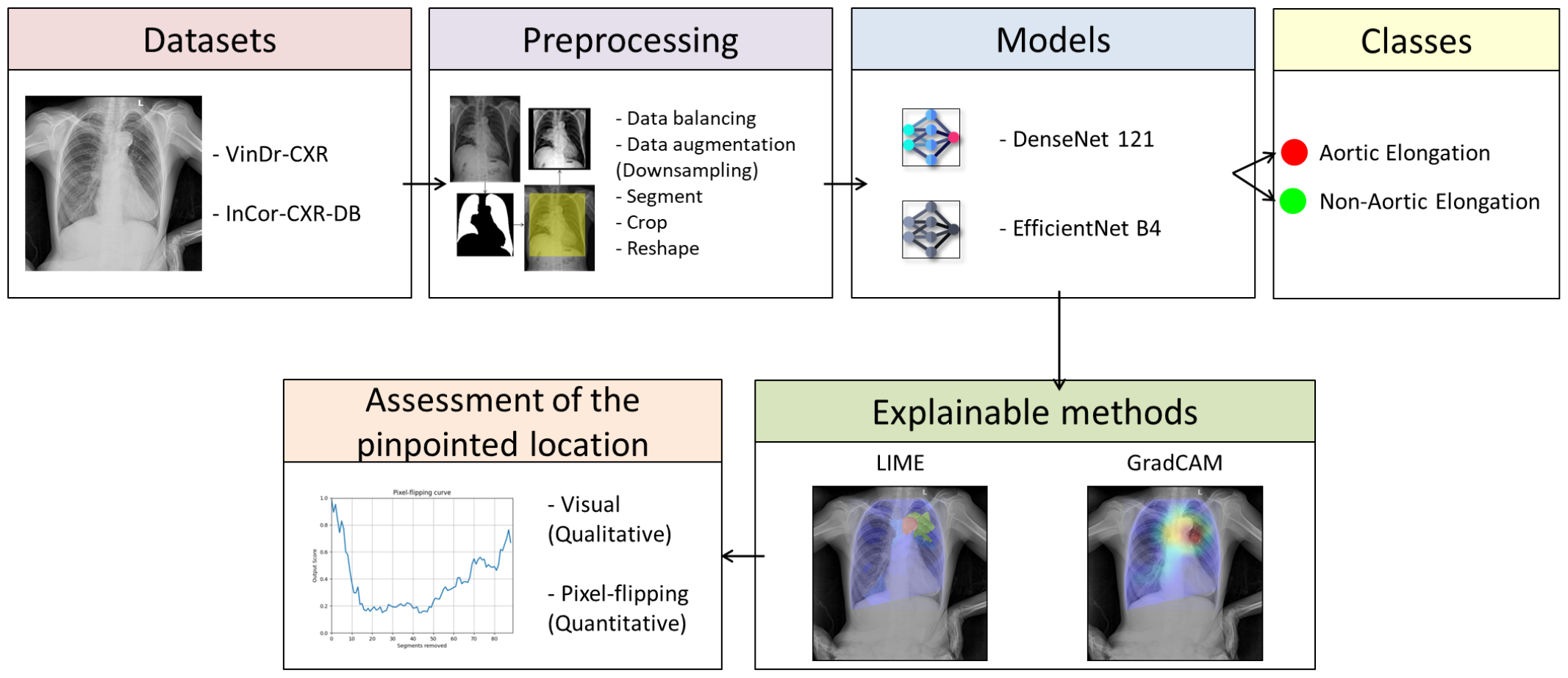
General structure of the proposed methodology.

### 2.1 Datasets

VinDr-CXR [17, 18, 21] is a dataset of CXR scans obtained retrospectively from two major hospitals in Vietnam. All images are in DICOM format and in postero-anterior (PA) view. It contains 18,000 images manually annotated by a group of radiologists. This dataset is divided into a training set with 15,000 scans independently labeled by three radiologists, and a test set with 3,000 scans labeled by the consensus of five radiologists. From these 15,000 exams, 10,606 are labeled as “Normal” and 4,376 have some abnormal condition, where 2,350 exams are labeled as Aortic Elongation. We downsampled this “Normal” class by randomly selecting 4,376 normal exams. Therefore, our resulting dataset consists of 8,752 exams. It should be stressed that due to the lack of patients ID information, images from the same patient may exist throughout train and test sets.

Besides the VinDr-CXR dataset, we used a private CXR dataset (InCor-CXR) collected from the Picture Archiving and Communication System (PACS) of a tertiary referral hospital in Brazil specialized in cardiology (Heart Institute Hospital), with patients older than 18 years old. All CXR images are in DICOM format and in a postero-anterior (PA) view. The text-based chest radiography reports were assessed by 2 experienced observers. This private dataset contains 473 exams labeled as Aortic Elongation and 760 exams labeled as non-Aortic Elongation, including equally sampled exams labeled as normal and abnormal conditions. The InCor-CXR dataset complies with all relevant ethical regulations and was approved by the Research Ethics Committee. Table 1 summarizes the information regarding both datasets.

**Table 1:** Number of CRX exams labeled as Aortic Elongation from VinDr-CXR dataset and our InCor-CRX private database.

|  | Datasets |  |
| --- | --- | --- |
|  | VinDr-CXR | InCor-CXR |
| Aortic Elongation | 2,350 | 473 |
| Non-Aortic Elongation | 6,402 | 760 |
| *Total* | *8,752* | *1,233* |

### 2.2 Image Preprocessing

To address the issue of the imbalanced data, we employed a downsampling technique on the larger class by randomly selecting exams labeled as non-aortic elongation. Subsequently, we applied a data augmentation technique to compensate for the reduced number of samples. Our preprocessing steps are based on previous works [7, 11] for cardiomegaly classification. In order to improve the efficiency of the training step and reduce the computational cost, we used a previously developed and validated model [20] that segments the lungs, to be able to create a binary mask of the chest cavity region, using a fine-tuned UNet-based CNN. We used the extreme points of this lung mask to create a bounding box and crop the chest cavity, segmenting only this region of interest that was used as input in our classification model. We rescaled the intensity of the cropped image with a contrast stretching method, including all intensities that fall within 1st and 99th percentiles of the image histogram. The image is converted into a square format through zero padding, ensuring the preservation of the tissue’s morphology depicted in the image, since in general, the networks expect square input images. Figure 2 displays the preprocessing steps described.

**Figure 2:**
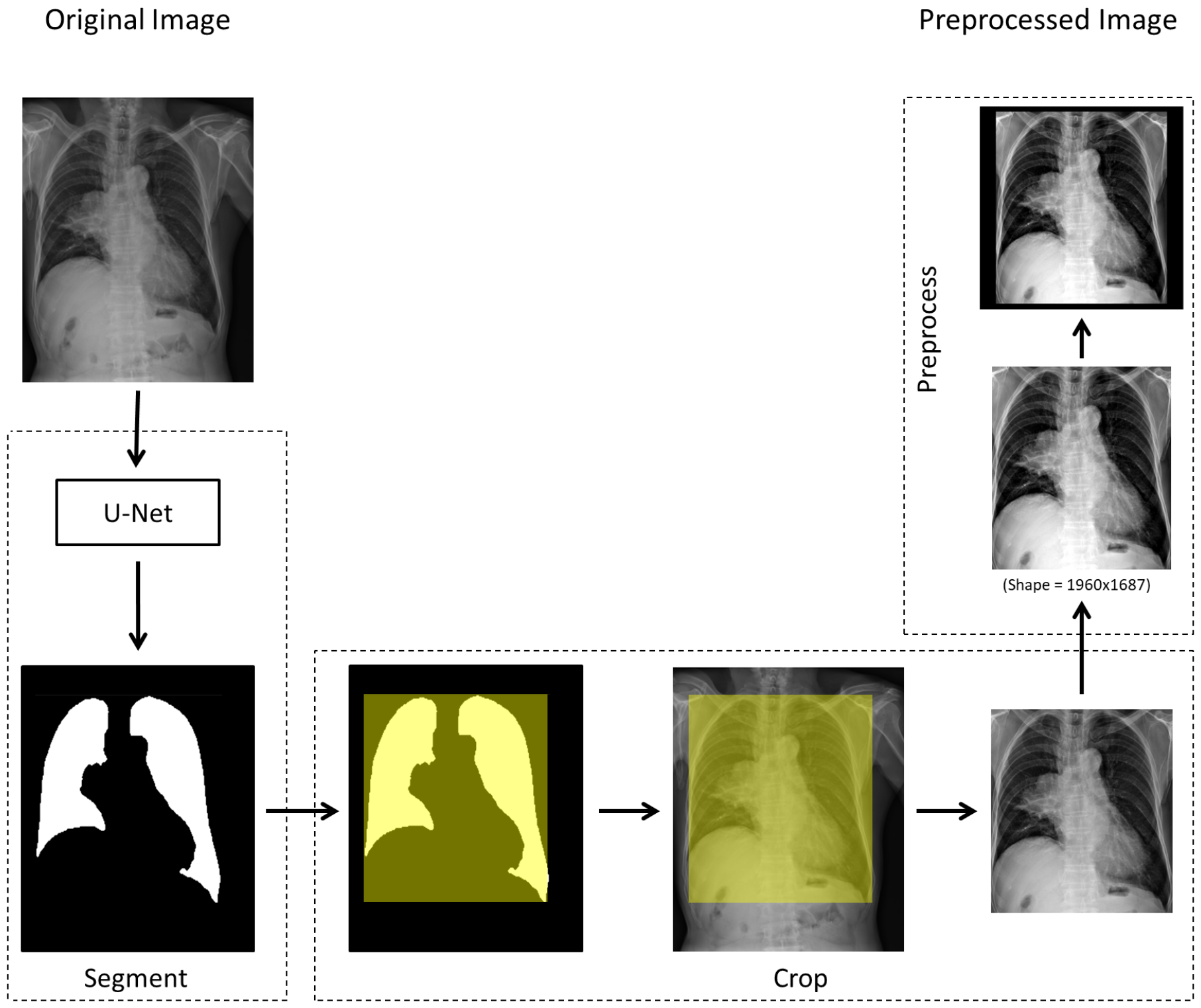
Steps for the proposed preprocessing task, being them: (i) chest cavity segmentation; (ii) cropping; (iii) contrast stretching; (iv) fill borders to get squared images; and (v) resize.

### 2.3 Deep Learning model

The small size of datasets in DL for analyzing medical images is a major constraint. Consequently, it is frequently difficult to train a CNN from scratch [4]. In this case, Transfer-learning is one solution. To do so, one can use a pre-trained network as a feature extractor and retrained it with the data from the new domain, such as CXR images. Here, we initiated the training with the ImageNet weights [25]. Then, we trained the models using the VinDr-CXR dataset to learn the patterns specific to the CXR domain. Subsequently, we performed fine-tuning using our InCor-CXR private dataset. Only the fully connected layers of the model were unfrozen during this fine-tuning process. Figure 3 depicts the overall proposed model based on transfer learning and fine-tunning techniques.

**Figure 3:**
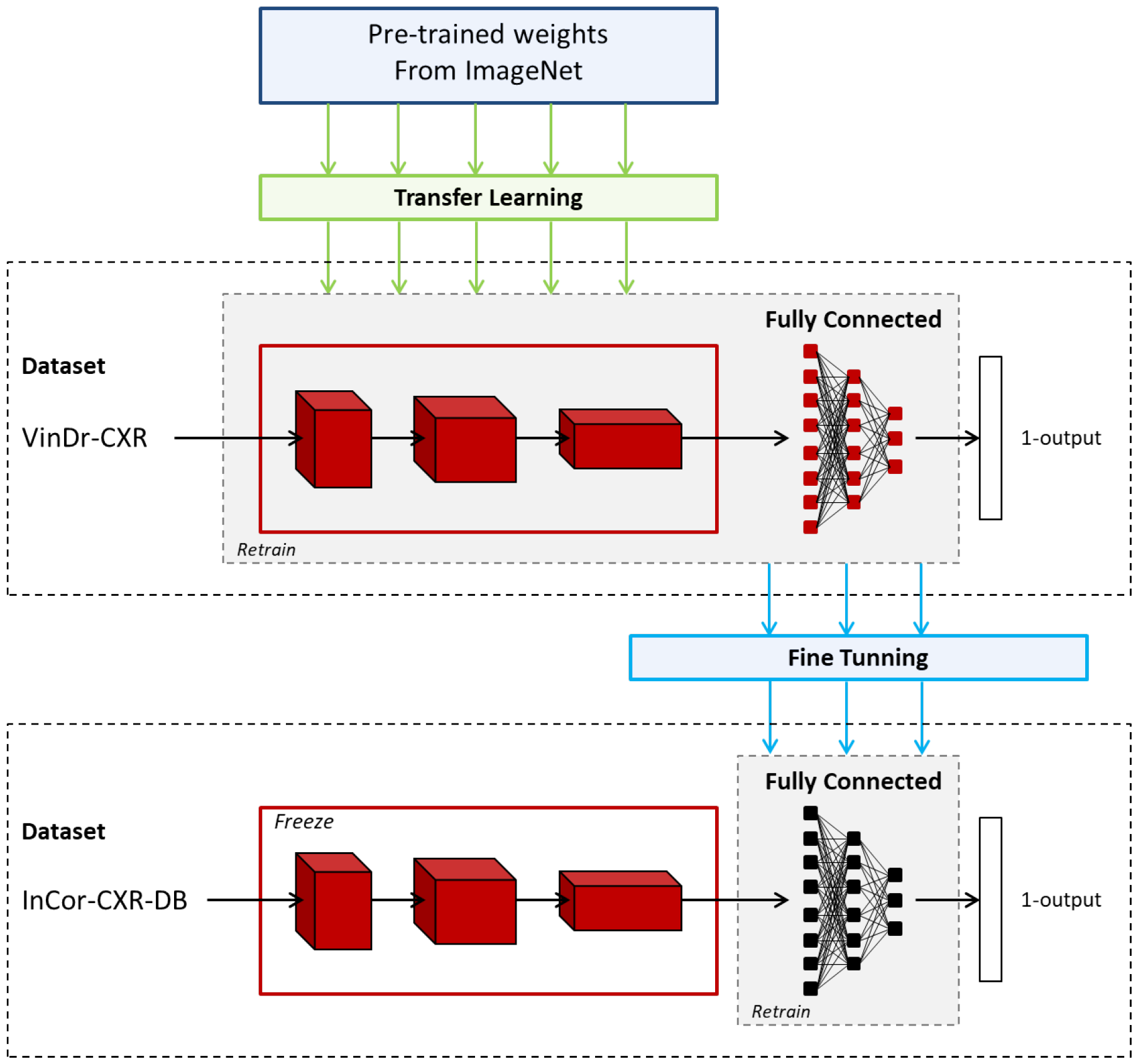
The transfer learning and fine-tuning techniques used in the development of the proposed CNN models.

For our experiments, we choose two different Deep Neural Network architectures that are widely used on the classification of CXRs, being them the DenseNet 121 and the EfficientNet B4 architectures. We used the default Keras architecture topology for the networks with an input resolution of 384×384×3 for the DenseNet 121 model, and an input resolution of 380x380x3 for the EfficientNet B4 model. For the fully connected layers, we used a customized 2-layer perceptron with dropout regularization of 30%, ReLU activation in the first layer (n = 256), and a sigmoid function in the last layer. Our model was trained over 40 epochs, with a 16 batch size per step using Adam optimizer with a learning rate of 0.0001 and a callback to reduce it by a factor of 0.2 every six epochs in case of no improvement in validation loss. Moreover, the data were augmented via: (1) rotation, (2) horizontal flipping, and vertical flipping. Our data augmentation strategy involved applying a 2× augmentation factor to the major class and an appropriately computed factor to the minor class. The purpose of this approach was to balance the representation of classes and address any potential class imbalance issues.

We implemented a robust 5-fold cross-validation strategy during the fine-tuning step of our model development process.

In order to evaluate the employed model, we assessed five different metrics, including Accuracy (Acc), Precision (Prec), Sensitivity (Se), Specificity (Spe), F1-score (F1), and Area Under Operational Receipt Curve (AUROC).

Moreover, first we utilized a cross-validation approach to estimate the performance of our models, which involved splitting our data into multiple subsets to train and evaluate the models iteratively. Once we obtained an understanding of their performance, we proceeded to retrain the models using the entire dataset, without the cross-validation process. This allowed us to create complete models which were subsequently applied to perform the proposed explainable AI methods.

Our experiment was performed using a Foxconn High-Performance Computer (HPC) M100-NHI with an 8 GPU cluster of 16 GB NVIDIA Tesla V100 cards. The methodology was implemented using the Python framework and Keras v2.4.3 with TensorFlow backend v2.3.0.

### 2.4 Explainable AI methods

We chose to employ two commonly used local explainable methods to analyze individual CXR images. These methods, LIME (Local Interpretable Model-agnostic Explanations) [22] and Grad-CAM (Gradient-weighted Class Activation Mapping) [28], are anticipated to offer valuable insights into the decision-making mechanisms of intricate machine learning models. The outcome of these analyses is visualized in a map where the most important regions are highlighted in red for both Grad-CAM and LIME methods.

LIME [22] is a model-agnostic method that explains the predictions of a machine learning model in a way that is expected to be easily interpretable to humans. LIME works by creating a local linear model around a specific data point to approximate how the original model makes its predictions. It then identifies the features, or superpixels (segments on the image instance) in the context of image inputs, of the input that are most important in determining the outcome of the prediction. In our work, we used a linear regression as the surrogate model, and segmented the images with a *quickshift* segmentation method from the skimage segmentation library on python, using the following parameters: kernel_size = 5; max_dist = 100; and ratio = 0.5. Moreover, we used 1000 perturbations to perform LIME.

On the other hand, Grad-CAM [28] is a visualization method that helps in understanding the regions of an image that contribute to the prediction made by a CNN. It uses the gradients of the predicted class with respect to the feature maps of the last convolutional layer to generate a heatmap that highlights the regions of the input image that are most relevant to the final prediction.

Both LIME and Grad-CAM are important tools for explainable AI as they help in identifying the features or regions of the input that the model is relying on to make its predictions. These methods can be used to identify potential errors in the model, identify biases, or simply to provide insights into how the model is processing the input data. By providing a more transparent and interpretable view into the inner workings of machine learning models, LIME and Grad-CAM help in building more trustworthy and reliable AI systems.

### 2.5 Quantification of Explainable AI methods

The pixel-flipping method [26, 27] is a technique used to quantify the explainability of AI methods that operate on image data, particularly in the context of computer vision tasks such as image classification. The goal of this method is to assess how sensitive the AI model’s predictions are to changes in individual pixels of an input image, which can provide insights into how the model makes decisions and which image regions are important for its predictions. The pixel-flipping method typically involves the following steps:

1. Input Image Selection: A set of input images from the InCor-CXR dataset used to test the AI model are selected for analysis. These images should represent the task of interest and cover a range of different samples to ensure a comprehensive assessment. Here we selected a subset of the InCor-CXR test set with 70 CXR exams labeled as Aortic Elongation;
2. Perturbation: Each selected image is then perturbed by flipping the value of individual pixels, either by inverting their intensity (e.g., changing a pixel from black to white or vice versa) or by changing their color value (e.g., changing a pixel from red to green or vice versa). In this work we set a pixel to zero for Grad-CAM method, or set a segment to zero for LIME method. This perturbation is done based on their importance as determined by the explainable methods, ordered from the most relevant to the least according to the particular explanation, showing how quickly the prediction score decreases. Likewise, for a baseline, we randomly selected the pixels or segments for comparison;
3. Prediction Evaluation: The perturbed images are then fed into the DL model, and the resulting predictions are compared to the original predictions on the unperturbed image. The discrepancy between the original and perturbed predictions is used as a measure of the model’s sensitivity to pixel-level changes. For example, if flipping a certain pixel significantly changes the model’s prediction, it indicates that the model may rely heavily on that pixel for its decision-making process. In this work we present the curves of the DL model output score for the perturbed images;
4. Interpretation: Finally, the results of the pixel-flipping analysis can be interpreted to gain insights into the model’s behavior. For example, if certain pixels have a high impact on the model’s predictions, it may indicate that those pixels are important for the model’s decision-making process and should be further investigated. This information can help understand the inner workings of the DL model and identify potential biases or limitations in its decision-making process.

Overall, the pixel-flipping method is a useful tool for quantifying the explainability of AI methods in computer vision tasks by assessing their sensitivity to pixel-level changes. Here, for LIME we gradually removed (set a segment to zero) the segments of an individual input image, ordered from the most relevant to the least according to the particular explanation, showing how quickly the prediction score decreases. For Grad-CAM, we removed 10, 20, …, 100% of the pixels, based on the most relevant on the resulting heatmap. Pixel-flipping curves are computed for a subset of the test set of individual images, and then averaged to create a mean curve. Finally, for comparison (baseline), we performed pixel-flipping by randomly removing the segments and pixels for LIME and Grad-CAM methods.

## 3 Results

The performance metrics of our models are outlined in Table 2. It can be observed that the DenseNet model achieved better results than the EfficientNet model in identifying Aortic Elongation. These results demonstrate the effectiveness of our approach in developing a reliable model for Aortic Elongation detection, which can potentially have significant implications for clinical practice.

**Table 2:** Performance of our proposed models for the identification of Aortic Elongation on CXR images.

| Model | Acc | Prec | Se (TPR) | Spe (TNR) | F1 | AUROC |
| --- | --- | --- | --- | --- | --- | --- |
| *EfficientNet B4* | 0.830 (0.009) | 0.771 (0.020) | 0.792 (0.014) | 0.854 (0.017) | 0.781 (0.009) | 0.909 (0.007) |
| *DenseNet 121* | 0.847 (0.007) | 0.756 (0.013) | 0.887 (0.027) | 0.823 (0.016) | 0.816 (0.010) | 0.931 (0.004) |

Figure 4 displays the results of the Grad-CAM and LIME methods for an exam labeled as Aortic Elongation (DenseNet prediction probability: 99.44%, EfficientNet prediction probability: 99.64%). Both explainable methods correctly identified the location of the aortic elongation.

**Figure 4:**
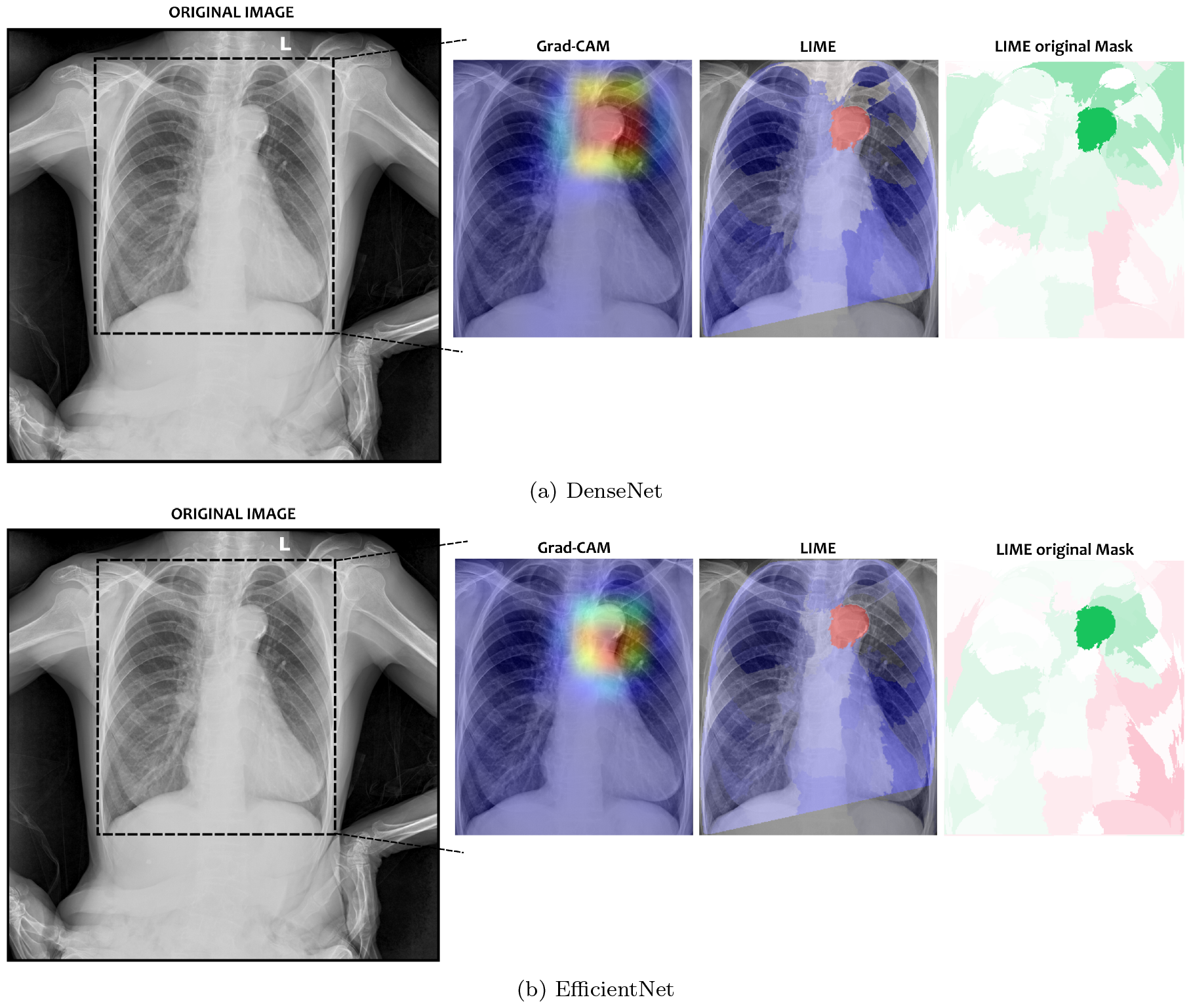
Explainable methods results for an exam correctly labeled as Aortic Elongation for both models. DenseNet prediction probability of aortic elongation: 99.44%. EfficientNet prediction probability of aortic elongation: 99.64%. In the original image, a region of interest is demarcated by a dashed box. The output of each method, Grad-CAM and LIME, is superimposed on the chest X-ray (CXR) image. The most relevant regions are highlighted in red, while the least relevant regions are indicated in blue. The “LIME original Mask” refers to the mask generated by the LIME method.

Similarly, Figure 5 shows the results of the explainable methods for an exam labeled as non-Aortic Elongation (DenseNet prediction probability: 0.60%, Efficient-Net prediction probability: 3.06%). Neither method highlighted any specific region since this is a normal CXR exam. For the DenseNet model, LIME highlighted some edge locations, while the Grad-CAM highlighted the lower right region of the thorax. In the case of the EfficientNet model, LIME highlighted various places in the lung region, while the Grad-CAM highlighted regions without the heart.

**Figure 5:**
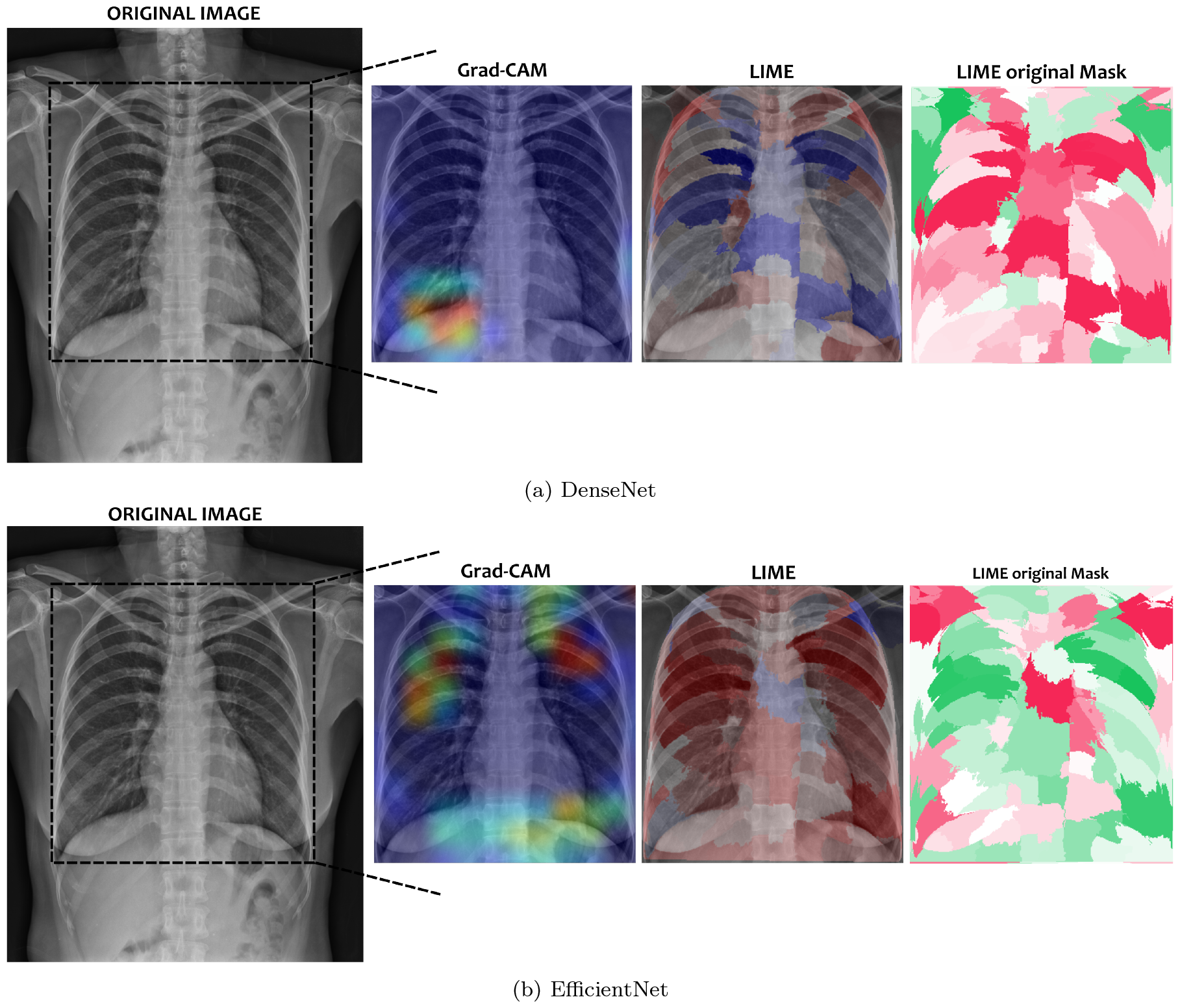
Explainable methods results for an exam correctly labeled as non-Aortic Elongation. DenseNet prediction probability of aortic elongation: 0.60%. EfficientNet prediction probability of aortic elongation: 3.06%. In the original image, a region of interest is demarcated by a dashed box. The output of each method, Grad-CAM and LIME, is superimposed on the chest X-ray (CXR) image. The most relevant regions are highlighted in red, while the least relevant regions are indicated in blue. The “LIME original Mask” refers to the mask generated by the LIME method.

Figure 6 presents the results of an exam that was wrongly predicted as Aortic Elongation but labeled as cardiomegaly in the dataset (DenseNet prediction probability: 80.16%, EfficientNet prediction probability: 92.81%). Here, all explainable methods identified the aorta region, except for the LIME method on the EfficientNet model, which highlighted the aorta region along with other parts of the chest, indicating the upper right region (near the shoulder) as the most important segment.

**Figure 6:**
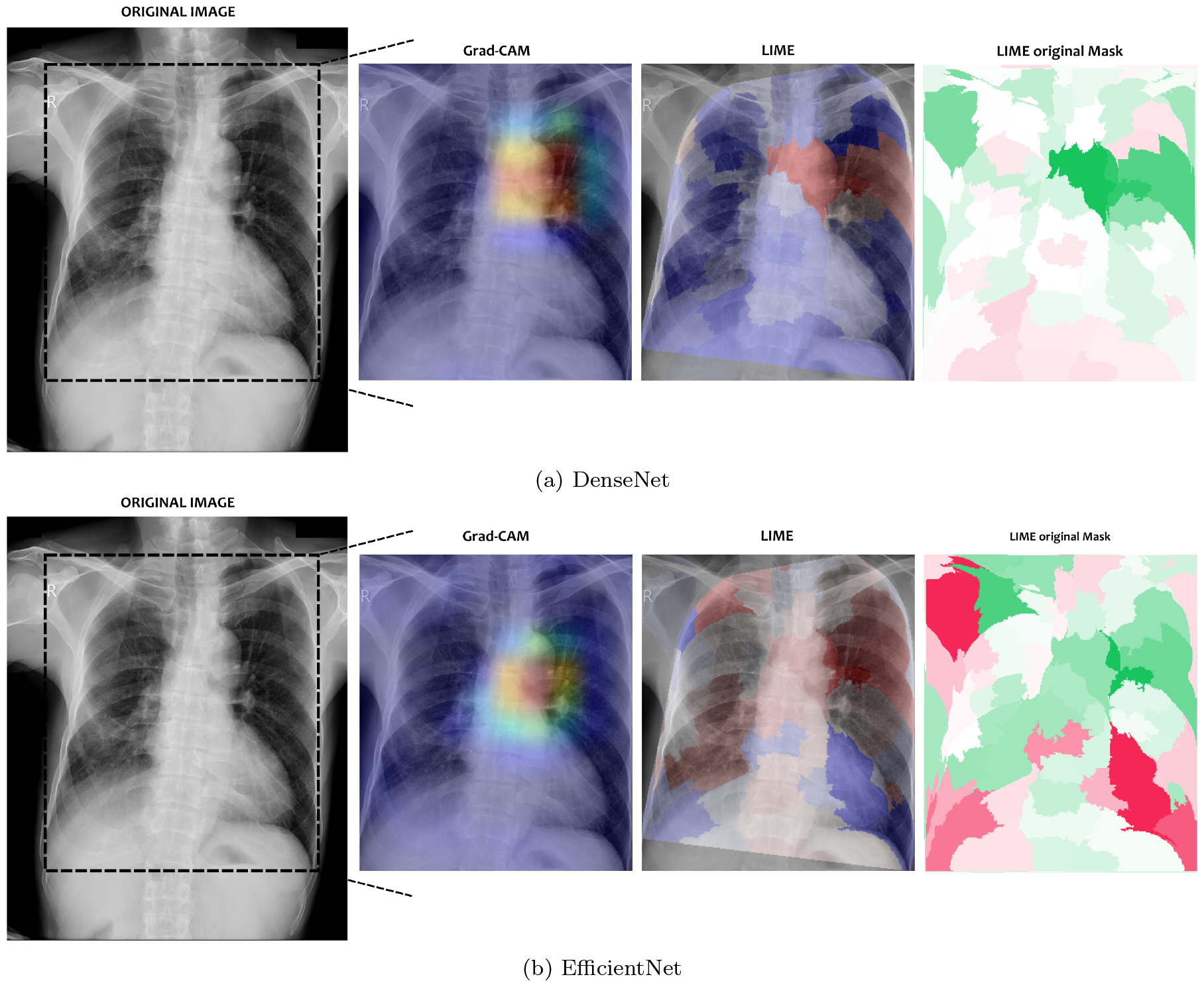
Explainable methods results for an exam wrongly labeled as Aortic Elongation. DenseNet prediction probability of aortic elongation: 80.16%. EfficientNet prediction probability of aortic elongation: 92.81%. In the original image, a region of interest is demarcated by a dashed box. The output of each method, Grad-CAM and LIME, is superimposed on the chest X-ray (CXR) image. The most relevant regions are highlighted in red, while the least relevant regions are indicated in blue. The “LIME original Mask” refers to the mask generated by the LIME method.

To assess the reliability of our studied explainable methods, we utilized the pixel-flipping method. The results of this quantitative method for our classification models are illustrated in Figure 7. There are significant differences in the results of both models. When relevant pixels/segments are removed, the DenseNet model experiences a rapid decline in its prediction performance (output score) for both Grad-CAM and LIME methods, which aligns with the desired behavior. However, for the EfficientNet model, the pixel-flipping method didn’t distinctly differentiate from the baseline of random removal.

**Figure 7:**
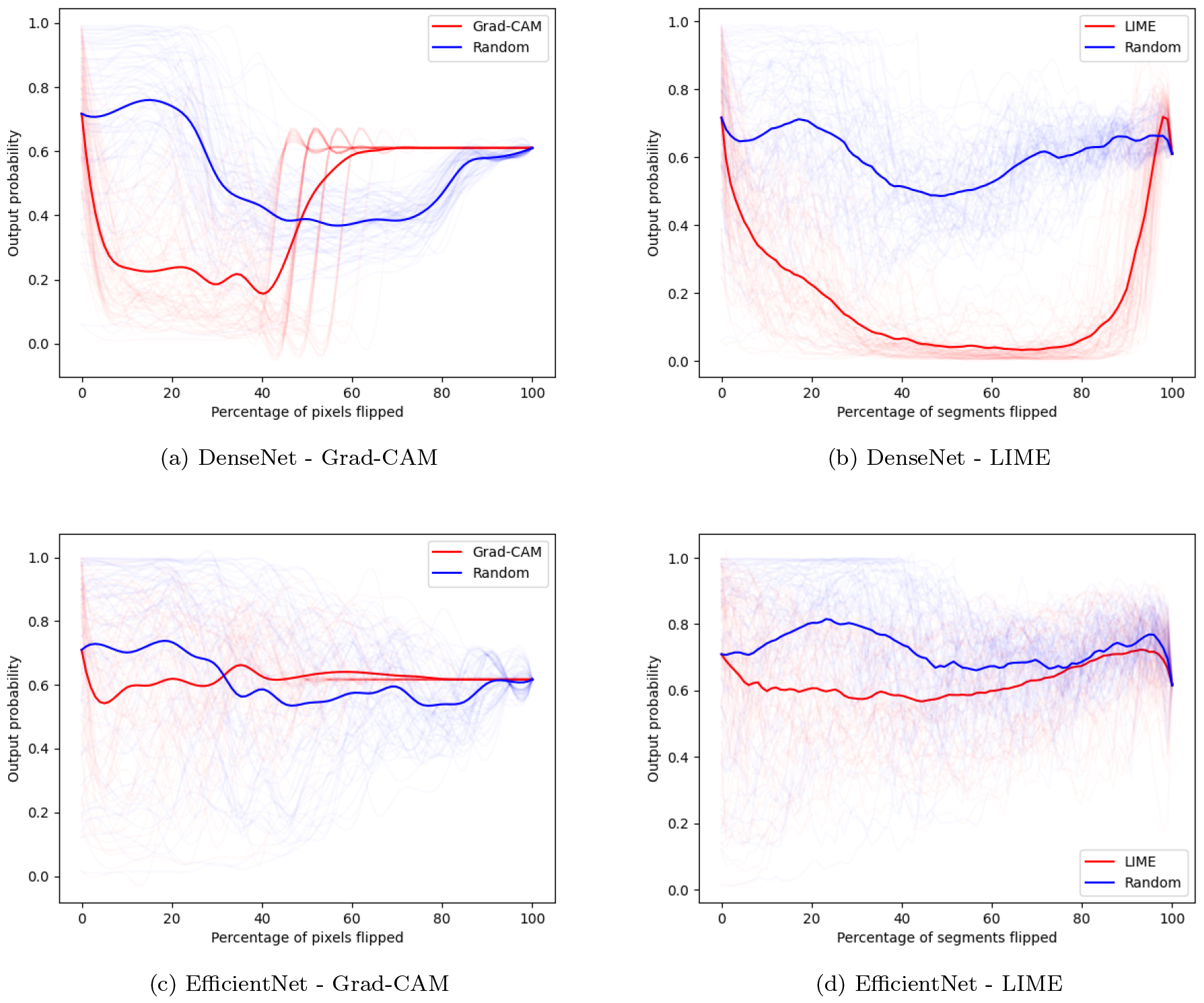
Pixel-flipping curves for a subset of individual CXR exams (thin lines) and their average mean curves (thick line) for both explainable methods, gradually removing the segments ordered from the most relevant (red lines) or randomly removing the segments (blue lines).

## 4 Discussion

We have developed a comprehensive strategy, by joining the Aortic Elongation model with the explainable methods, to analyze Chest X-rays. This approach has the potential to enhance diagnostic accuracy and aid in early detection of this cardiac condition. Through transfer learning and fine-tunning processes, our models were able to accurately identify Aortic Elongation. By using explainable methods, we can provide valuable insights for physicians into the decision making process of our models. Chest X-rays are often considered a cost-effective and time-efficient option, making them an ideal initial screening tool. By utilizing this approach, high-risk patients can be efficiently triaged during their routine care. Subsequently, appropriate lifestyle modification counseling can be provided, and further examinations or interventions can be pursued if necessary.

In a qualitative analysis, Grad-CAM and LIME explainable methods correctly pinpointed the expected aorta region on exams correctly predicted as Aortic Elongation. On the first example (Figure 4), it is interesting to observe that both models presented different explanations. One could argue that different models learn the problem (Aortic Elongation prediction, here) differently, and the visual explanations could assess if they learned the problem correctly. On the second example (Figure 5), we can see that for this normal CXR the models seems to not know where to look on the image. One could suggest that the model didn’t find the aorta problem, making the explainable methods highlighting spurious regions of the image. Moreover, again we can see the difference between both models, where DenseNet only highlights few segments, whereas EfficientNet indicate more regions on the CXR. In the case where the models wrongly predicted Aortic Elongation (Figure 6) both models indicated the aorta region as the most relevant.

Quantitatively, it is expected that the explainable methods which accurately highlight the image pixels/segments influencing the model’s decision will exhibit a rapid decline in prediction performance (mean output score) as these pixels/segments are progressively perturbed (set to zero). To establish a baseline, random explanations (blue lines) are also included on Figure 7, as it is anticipated that perturbing image pixels/segments randomly will result in a slower decrease in performance. In addition, it is important to clarify that the results obtained through the pixel-flipping method reflect the output score of the models and not their accuracy. The output score represents the confidence or certainty assigned by the model to a specific prediction. It is distinct from accuracy, which measures the overall correctness of the model’s predictions.

Based on our findings presented in Figure 7, several observations can be highlighted: (i) The DenseNet model demonstrated the expected behavior for both explainable methods; (ii) Grad-CAM applied to the DenseNet model showed a rapid decrease in prediction performance. When 50% of the pixels were removed, the prediction performance reached a basal output score level on average; (iii) LIME performed better than Grad-CAM on the DenseNet model and significantly distinguished itself from the baseline (random removal). Removing segments rapidly led to a drop in prediction performance. Removing the most important segments made the model to change prediction to non-Aortic Elongation. However, with the removal of the final segments, the performance reached the same basal level as aforementioned for the Grad-CAM; (iv) The EfficientNet model did not clearly differentiate itself from the baseline (random removal) for both explainable methods; and (v) Both models concluded with a basal output score prediction of 0.61. This suggests that if we take an CXR image and set all pixels/segments to zero, the model, on average, would classify it as Aortic Elongation using a threshold of 0.5. However, it’s important to note that a completely black image (with all pixels set to zero) falls outside the domain on which the model was trained. Therefore, the model is not fitted to recognize this pattern since it hasn’t learned it. These observations provide valuable insights into the behavior and performance of the DenseNet and EfficientNet models when subjected to the explainable and pixel-flipping methods.

To the best of our knowledge, there are only a limited number of studies that have focused on the automated detection of Aortic Elongation specifically from chest X-rays. It is important to note that some studies employ a multilabel paradigm, where Aortic Elongation may be included as one of the labels depending on the dataset. However, the number of works specifically targeting the detection of Aortic Elongation in chest X-rays remains relatively scarce. Rosenwasser et al. (2022) [23] used an Inception V3 and a DenseNet169 models on the VinDr-CXR dataset, relating this cardiac condition with Marfan’s syndrome. Likewise, Lee et al. (2022) [13] propose the detection of aortic dissection based on a ResNet 18 CNN, using Grad-CAM to visually interpret their results. Hence, it is important to emphasize the significance of automating the identification of these conditions, given their potential implications for patient health and the effectiveness of medical interventions.

Many remote hospitals lack access to radiologists, making computer-aided diagnostic systems crucial for accurate results. These systems aid inexperienced doctors, reducing expenses, prioritizing critical cases, and shortening wait times. However, their adoption in clinical practice is rare due to: (i) inconsistencies in existing datasets extracted from radiology reports using NLP methods; (ii) scarcity of labeled CXR datasets; (iii) research focusing more on model performance than practical application; and (iv) complexity of machine learning algorithms hindering interpretability.

It is indispensable that the models manage to generate explanations about their decision making. The issue of interpretability cannot be a limiting factor for systems already developed. We must make use of the great existing capacity of AI systemsto generate more efficient and reliable models. Grad-CAM and LIME methods can provide valuable insights into how AI models make decisions and identify important image regions for their predictions, which can be useful for improving model transparency, fairness, and trustworthiness. Moreover, Pixel-flipping can give a quantitative metric to the explainable methods, allowing assessing their interpretations.

Taken together, our results demonstrate the effectiveness of our approach in developing a reliable model for Aortic Elongation detection. The high accuracy, precision, and AUROC scores achieved by the DenseNet model, along with the supporting visualizations from the explainable methods, underscore its potential value in clinical practice. Further research and development in this area could lead to improved diagnostic capabilities and enhance patient care in the field of cardiac imaging.

## 5 Conclusion

Our study presents a strategy for analyzing CXR to detect Aortic Elongation through DL models and XAI methods. Our approach demonstrates promising results in enhancing diagnostic accuracy and aiding early detection. By leveraging transfer learning techniques, our models accurately identify Aortic Elongation, with explainable methods providing valuable insights into their decision-making processes for physicians. Our findings highlight the potential of AI-driven diagnostic systems to streamline healthcare processes and prioritize patient care. Continued research in this area is important for further advancements in cardiac imaging and the broader adoption of AI technologies in clinical practice.

## Data Availability

Publicly available datasets were analyzed in this study. Private InCor-CXR-DB dataset will be made available upon reasonable request to the authors.

## Acknowledgements

This study was supported by São Paulo Research Foundation (FAPESP) – grant n^*o*^ 2021/12935-0, the Zerbini Foundation, and Foxconn Brazil, as part of the research project “Machine Learning in Cardiovascular Medicine”.

## Ethics Statement

This research was approved by the Internal Review Board (IRB), registration CAAE 45070821.3.0000.0068, as part of the Machine Learning in Cardiovascular Medicine Project.

## Author contributions statement

E.R. Conceptualization, Methodology, Implementation and Writing. D.A.C.C. Conceptualization, Methodology, Implementation and Writing. F.M.D. Methodology and Writing. J.E.K. and M.A.G. acted as project leaders. All authors analyzed the results and revised critically the manuscript. All authors read and approved the submitted manuscript.

## Competing interests

The authors declare no competing interests.

## Notes

### Competing Interest Statement

The authors have declared no competing interest.

### Funding Statement

This study was supported by Sao Paulo Research Foundation (FAPESP) grant no 2021/12935-0, the Zerbini Foundation, and Foxconn Brazil, as part of the research project Machine Learning in Cardiovascular Medicine.

### Author Declarations

This research was approved by the Internal Review Board (IRB) of the Heart Institute (InCor) Clinics Hospital University of Sao Paulo Medical School (HCFMUSP), registration CAAE 45070821.3.0000.0068, as part of the Machine Learning in Cardiovascular Medicine Project.

### Summary of Updates

We revised the text, changed the template and added a conclusion section.

## References

[1] A decade of discovery in the genetic understanding of thoracic aortic disease. Canadian Journal of Cardiology, 32(1):13–25, 2016.

[2] Bouke P Adriaans, Samuel Heuts, Suzanne Gerretsen, Emile C Cheriex, Rein Vos, Ehsan Natour, Jos G Maessen, Peyman Sardari Nia, Harry J G M Crijns, Joachim E Wildberger, and Simon Schalla. Aortic elongation part i: the normal aortic ageing process. Heart, 104(21):1772–1777, 2018.

[3] Koichi Akutsu, Atsushi Watanabe, Takeshi Yamada, Tomoko Sahara, Sayuri Hiraoka, and Wataru Shimizu. Vascular involvements are common in the branch arteries of the abdominal aorta rather than in the aorta in vascular ehlers-danlos syndrome. CJC Open, 5(1):72–76, 2023.

[4] Ivo M. Baltruschat, Hannes Nickisch, Michael Grass, Tobias Knopp, and Axel Saalbach. Comparison of Deep Learning Approaches for Multi-Label Chest X-Ray Classification. ScientificReports, 9(1):6381, 2019.

[5] Carolyn A Bondy. Aortic dissection in turner syndrome. CurrentOpinioninCardiology, 23(6):519– 526, 2008.

[6] Bert Callewaert, Fransiska Malfait, Bart Loeys, and Anne De Paepe. Ehlers-danlos syndromes and marfan syndrome. BestPractice/ResearchClinicalRheumatology, 22(1):165–189, 2008. Orphan Skeletal Diseases.

[7] Diego Cardenas, José Ferreira Junior, Ramon Moreno, Marina Rebelo, José Krieger, and Marco Gutierrez. Multicenter validation of convolutional neural networks for automated detection of cardiomegaly on chest radiographs. In AnaisdoXXSimpósioBrasileirodeComputaçãoAplicadaàSaúde, pages 179–190, Porto Alegre, RS, Brasil, 2020. SBC.

[8] Marzyeh Ghassemi, Luke Oakden-Rayner, and Andrew L. Beam. The false hope of current approaches to explainable artificial intelligence n health care. LancetDigitalHealth, 3(11):e745–e750, 2021.

[9] Judith Z. Goldfinger, Jonathan L. Halperin, Michael L. Marin, Allan S. Stewart, Kim A. Eagle, and Valentin Fuster. Thoracic aortic aneurysm and dissection. JournaloftheAmericanCollegeofCardiology, 64(16):1725–1739, 2014.

[10] Alan Graham Stuart and Andrew Williams. Marfan’s syndrome and the heart. Archives of Disease in Childhood, 92(4):351–356, 2007.

[11] José Raniery Ferreira Junior, Diego Armando Cardona Cardenas, Ramon Alfredo Moreno, Marina de Fátima de Sá Rebelo, José Eduardo Krieger, and Marco Antonio Gutierrez. A general fully automated deep-learning method to detect cardiomegaly in chest x-rays. In Maciej A. Mazurowski and Karen Drukker, editors, Medical Imaging2021:Computer-AidedDiagnosis, olume 11597, pages 537 – 542. International Society for Optics and Photonics, SPIE, 2021.

[12] Shinjini Kundu. AI in medicine must be explainable. NatureMedicine, 27(8):1328–1328, 2021.

[13] Dong Keon Lee, Jin Hyuk Kim, Jaehoon Oh, Tae Hyun Kim, Myeong Seong Yoon, Dong Jin Im, Jae Ho Chung, and Hayoung Byun. Detection of acute thoracic aortic dissection based on plain chest radiography and a residual neural network (resnet). Scientific Reports, 12:21884, 2022.

[14] Scott Lundberg and Su-In Lee. A unified approach to interpreting model predictions, 2017. arXiv: 1705.07874.

[15] Gretchen MacCarrick, James H. Black, Sarah Bowdin, Ismail El-Hamamsy, Pamela A. Frischmeyer-Guerrerio, Anthony L. Guerrerio, Paul D. Sponseller, Bart Loeys, and Harry C. Dietz. Loeys–dietz syndrome: a primer for diagnosis and management. Genetics in Medicine, 16(8):576–587, 2014.

[16] Christoph Molnar. Interpretable Machine Learning. 2019.

[17] Ha Q. Nguyen, Khanh Lam, Linh T. Le, Hieu H. Pham, Dat Q. Tran, Dung B. Nguyen, Dung D. Le, Chi M. Pham, Hang T. T. Tong, Diep H. Dinh, Cuong D. Do, Luu T. Doan, Cuong N. Nguyen, Binh T. Nguyen, Que V. Nguyen, Au D. Hoang, Hien N. Phan, Anh T. Nguyen, Phuong H. Ho, Dat T. Ngo, Nghia T. Nguyen, Nhan T. Nguyen, Minh Dao, and Van Vu. Vindr-cxr: An open dataset of chest x-rays with radiologist’s annotations, 2020.

[18] Ha Q. Nguyen, Khanh Lam, Linh T. Le, Hieu H. Pham, Dat Q. Tran, Dung B. Nguyen, Dung D. Le, Chi M. Pham, Hang T. T. Tong, Diep H. Dinh, Cuong D. Do, Luu T. Doan, Cuong N. Nguyen, Binh T. Nguyen, Que V. Nguyen, Au D. Hoang, Hien N. Phan, Anh T. Nguyen, Phuong H. Ho, Dat T. Ngo, Nghia T. Nguyen, Nhan T. Nguyen, Minh Dao, and Van Vu. Vindr-cxr: An open dataset of chest x-rays with radiologist’s annotations (version 1.0.0), 2020.

[19] Keiron O’Shea and Ryan Nash. An introduction to convolutional neural networks, 2015.

[20] I. Pazhitnykh and V. Petsiuk. Lung segmentation 2d, 2017.

[21] Hieu H. Pham, Ha Q. Nguyen, Hieu T. Nguyen, Linh T. Le, and Lam Khanh. An accurate and explainable deep learning system improves interobserver agreement in the interpretation of chest radiograph. IEEE Access, 10:104512–104531, 2022.

[22] Marco Tulio Ribeiro, Sameer Singh, and Carlos Guestrin. “why should i trust you?”: Explaining the predictions of any classifier, 2016. arXiv: 1602.04938.

[23] Tom Rosenwasser, Ronit Lain, and Miri Weiss Cohen. Aortic enlargement detection using chest x-rays to identify potential marfan syndrome. Procedia Computer Science, 207:2125–2133, 2022. Knowledge-Based and Intelligent Information Engineering Systems: Proceedings of the 26th International Conference KES2022.

[24] Cynthia Rudin. Stop explaining black box machine learning models for high stakes decisions and use interpretable models instead. NatureMachineIntelligence, 1(5):206–215, 2019.

[25] Olga Russakovsky, Jia Deng, Hao Su, Jonathan Krause, Sanjeev Satheesh, Sean Ma, Zhiheng Huang, Andrej Karpathy, Aditya Khosla, Michael Bernstein, Alexander C. Berg, and Li Fei-Fei. ImageNet Large Scale Visual Recognition Challenge. InternationalJournalofComputerVision, 115:211–252, 2015.

[26] Wojciech Samek, Alexander Binder, Grégoire Montavon, Sebastian Lapuschkin, and Klaus-Robert Müller. Evaluating the visualization of what a deep neural network has learned. IEEETransactionsonNeuralNetworksandLearningSystems, 28(11):2660–2673, 2017.

[27] Wojciech Samek, Grégoire Montavon, Sebastian Lapuschkin, Christopher J. Anders, and Klaus-Robert Müller. Explaining deep neural networks and beyond: A review of methods and applications. ProceedingsoftheIEEE, 109(3):247–278, 2021.

[28] Ramprasaath R. Selvaraju, Michael Cogswell, Abhishek Das, Ramakrishna Vedantam, Devi Parikh, and Dhruv Batra. Grad-cam: Visual explanations from deep networks via gradient-based localization. In 2017 IEEE International Conference on ComputerVision(ICCV), pages 618–626, 2017.

[29] Karen Simonyan, Andrea Vedaldi, and Andrew Zisserman. Deep inside convolutional networks: Visualising image classification models and saliency maps, 2014. arXiv: 1312.6034.

[30] Rikiya Yamashita, Mizuho Nishio, Richard Kinh Gian Do, and Kaori Togasho. Convolutional neural networks: An overview and application in radiology. Insightsintoimaging, 9:611–629, 2018.

